# Exploring the Application of Target Trial Emulation in Vaccine Evaluation: Scoping Review

**DOI:** 10.1101/2024.07.26.24311066

**Authors:** Toshiaki Komura, Miwa Watanabe, Kayoko Shioda

## Abstract

**Background:** Target trial emulation (TTE) has gained popularity in evaluating treatments and health interventions. Its application to infectious disease outcomes requires careful consideration, as infectious disease transmission violates the assumption of no interference. We conducted a scoping review to understand how TTE approaches have been applied to vaccine evaluation.

**Methods:** We conducted a systematic search of literature published in PubMed, Embase, and Web of Science until May 2024, using keywords related to TTE, infectious diseases, and vaccines. Three independent reviewers screened titles and abstracts for relevance. Full-text articles meeting inclusion criteria were further assessed for eligibility.

**Results:** Our keyword-based search and citation search identified a total of 240 studies. Of these, 34 original research studies used TTE approaches to evaluate vaccines, predominantly published from 2022 to 2024. Most studies (n=32, 94%) were conducted in high-income countries. The majority (n=31, 91%) evaluated the effect of COVID-19 vaccines, with one study each evaluating influenza, mpox, and rotavirus vaccines. Nationwide healthcare databases were used in 17 studies (50%). Twenty-one studies (62%) conducted analysis among adults aged ≥18 years, while six studies (18%) focused on children <18 years. Most studies did not define which of the four effects of vaccination they evaluated (direct, indirect, total, or overall effect), and none incorporated interference in vaccine evaluation.

**Conclusions:** Our review highlights the increasing popularity of TTE in vaccine evaluation following the COVID-19 pandemic. Further discussions are needed to establish TTE approaches to estimating four effects of vaccination, using large, routinely collected data.

## Key Messages

- This study performed a scoping review on how target trial emulation (TTE) approaches have been applied to vaccine evaluation, which requires careful consideration of interference.
- Among 34 studies in our scoping review, the majority evaluated the effect of COVID-19 vaccines, with one study each evaluating influenza, mpox, and rotavirus vaccines. Most studies were conducted in high-income countries, predominantly published from 2022 to 2024.
- Most studies did not define which of the four effects of vaccination they evaluated (direct, indirect, total, or overall effect), and none incorporated interference in vaccine evaluation.
- Our scoping review indicates a need for careful discussions to estimate the four effects of vaccination, using large, routinely collected data in the TTE framework.

## INTRODUCTION

Target trial emulation (TTE) is a causal inference framework that allows us to emulate a hypothetical target trial using observational data. TTE has become a popular research method to evaluate treatment and health interventions due to its practical and methodological advantages, serving as an important alternative to randomized controlled trials (RCTs) (1). While head-to-head RCTs are widely regarded as the gold standard for intervention evaluation, they are time-consuming, and operational and ethical challenges often hinder the randomization of participants in RCTs. Even when RCTs are feasible, they are generally restricted to small sample sizes and short follow-up periods, making it difficult to detect rare or long-term outcomes (2). RCTs also face constraints in comparing a large number of different protocols with various doses and treatment schedules due to resource limitations. Additionally, RCTs are primarily conducted in urban settings in high-income countries with selected population groups (3,4), leaving underrepresented groups reliant on findings from contexts that may not align with their own (5,6).

Observational studies could become useful to address these challenges posed by RCTs, especially when leveraging existing surveillance systems or large-scale health data (7). Existing surveillance systems can provide affordable and readily available data for intervention evaluation, helping us overcome operational and ethical challenges with RCTs. If surveillance systems cover the whole nation for a long time period, they could provide important information on short- and long-term outcomes of various diseases across demographic characteristics and geographic locations. However, as widely discussed, observational studies come with their own set of analytic challenges, including selection bias, confounding, and immortal time bias (which occurs when a time during which the outcome could not have happened is incorrectly handled in the analysis, often due to the misalignment of eligibility criteria and treatment assignment) (7–12).

TTE is a promising tool to harness the benefits of observational data while mitigating their analytic challenges (1). TTE facilitates causal inference using observational data, by mimicking a hypothetical target trial. By clearly specifying "time-zero" for treatment and control groups, TTE aims to effectively remove selection bias and immortal time bias. With the blurred lines between methodological advances in RCTs and observational studies, TTE has garnered attention as a convenient design for evaluating interventions (13).

While TTE has promising applications, it requires careful attention when used in the context of infectious diseases, where outcomes often do not satisfy the fundamental assumption of causal inference—no interference (14,15). Infectious diseases, which spread from infected individuals to susceptible individuals, challenge the assumption that the treatment of one person does not affect another person (16,17). This concern becomes particularly important when evaluating the effect of vaccines, because vaccination reduces population-level transmission, subsequently lowering the risk of infection among both vaccinated and unvaccinated individuals. In fact, vaccine evaluation has well-established guidelines categorizing vaccine effects into the following four types (18,19):

1. *direct effect*, which is the difference between the outcome in the vaccinated individual and what the outcome would have been without vaccination in the same person;
2. *indirect effect*, which is the difference in the outcome in the unvaccinated individual in a population with a vaccination program and what the outcome would have been in the unvaccinated individual in a population without a vaccination program;
3. *total effect,* which is the difference between the outcome in the vaccinated individual in a population with a vaccination program and what the outcome would have been in the unvaccinated individual in a comparable population without a vaccination program; and
4. *overall effect,* which is the difference in the outcome in an average vaccinated individual in a population with a vaccination program compared with the outcome of an unvaccinated individual in a comparable population without a vaccination program.

In light of these considerations, we explored how the TTE framework has been applied to vaccine evaluation. This scoping review investigated the recent trends in the use of TTE for assessing vaccine effectiveness and safety. We summarized the target populations, data sources, vaccine products, comparators, and clinical outcomes. Additionally, we examined specific TTE and statistical methods used in each study, and investigated how studies defined specific types of vaccine effects within the TTE framework. Finally, we evaluated whether and how studies addressed the dependent nature of infectious diseases. Based on findings from the scoping review, we highlighted methodological gaps and provided recommendations for future studies using TTE in the field of vaccine evaluation.

## METHODS

### Search strategy

We followed the Preferred Reporting Items for Systematic Reviews and Meta-Analyses (PRISMA) extension for a scoping review checklist to design this scoping review (20). The protocol of this scoping review was registered with the Open Science Framework Registries on January 29, 2024 (21). Studies were extracted from three databases: PubMed, Embase, and Web of Science. The databases were searched from January 1, 2012, following the protocol of a previous systematic review (22), to May 21, 2024. Studies published in English were included. The search term included keywords related to vaccine evaluation and target trial emulation and was determined through group consensus (**Supplementary Table 1**). As TTE approaches are relatively new, especially in the field of vaccine evaluation, studies may have not explicitly stated that TTE approaches were used. The authors aimed to identify these studies by going through the literature and citations of identified studies.

### Eligibility Criteria

Three independent reviewers (TK, MW, and KS) screened titles and abstracts based on the following inclusion criteria: 1) original research articles, 2) primary objectives that include the evaluation of vaccines, 3) study designs that adhere to a target trial emulation framework, and 4) peer-reviewed articles published in scientific journals.

### Data Extraction

To examine study characteristics, we extracted the following information: authors, year of publication, countries of origin, target populations, data sources, treatments (i.e., evaluated vaccine regimen), comparators, outcomes, types of vaccine effects evaluated, specific TTE methods, applied statistical methods, and whether the interference was adjusted, incorporated, or discussed. Three reviewers (TK, MW, and KS) independently conducted the full-text screening.

## RESULTS

### Characteristics, target populations, and data sources of included studies

A total of 234 studies were identified from the keyword-based literature search, and 28 of them met the inclusion criteria after the screening (**Figure 1**) (23–50). Additionally, we identified six studies through citation search that did not explicitly state that the TTE approach was used but applied it for vaccine evaluation (51–56). Therefore, a total of 34 studies were included in the analysis. Most studies were published from 2022 to 2024 (2021: n=5, 15%; 2022: n= 10, 29%; 2023: n= 12, 35%; and 2024: n=7, 21%) (**Table 1**). The United States (n= 14, 41%)(24,27–29,32,38–40,44,45,47,48,50,51) and the United Kingdom (n= 5, 15%)(26,30,36,37,43) were the most common countries represented in the studies. According to the World Bank classification (57), almost all studies (n=32, 94%) were conducted in high-income countries (23–33,35–48,50–56), except for two (6%) in upper-middle-income countries (34,49). Regarding the target population, nine studies (26%) conducted analysis among veterans (24,27–29,32,38,39,45,47), and one study (3%) was conducted among nursing home residents (40). Twenty-one studies (62%) conducted analysis among adults aged ≥18 years (24,26–30,32–34,36–39,41,43–49), three of which (9%) specifically focused on older adults aged ≥60 years (26,34,43). Six studies (18%) focused on children <18 years (23,31,35,51,55,56). Among studies conducted in the U.S., the most common data source was the national healthcare databases of the Department of Veterans Affairs (n= 9, 64%) (24,27–29,32,38,39,45,47), whereas, in other countries, national healthcare service data, primary care data, cohort data, surveillance data, vaccine registry data, and insurance service data were used. Nationwide healthcare databases were used in 17 studies (50%) (23–25,27–29,32,35,36,38,39,41,42,45–47,49).

**Figure 1.**
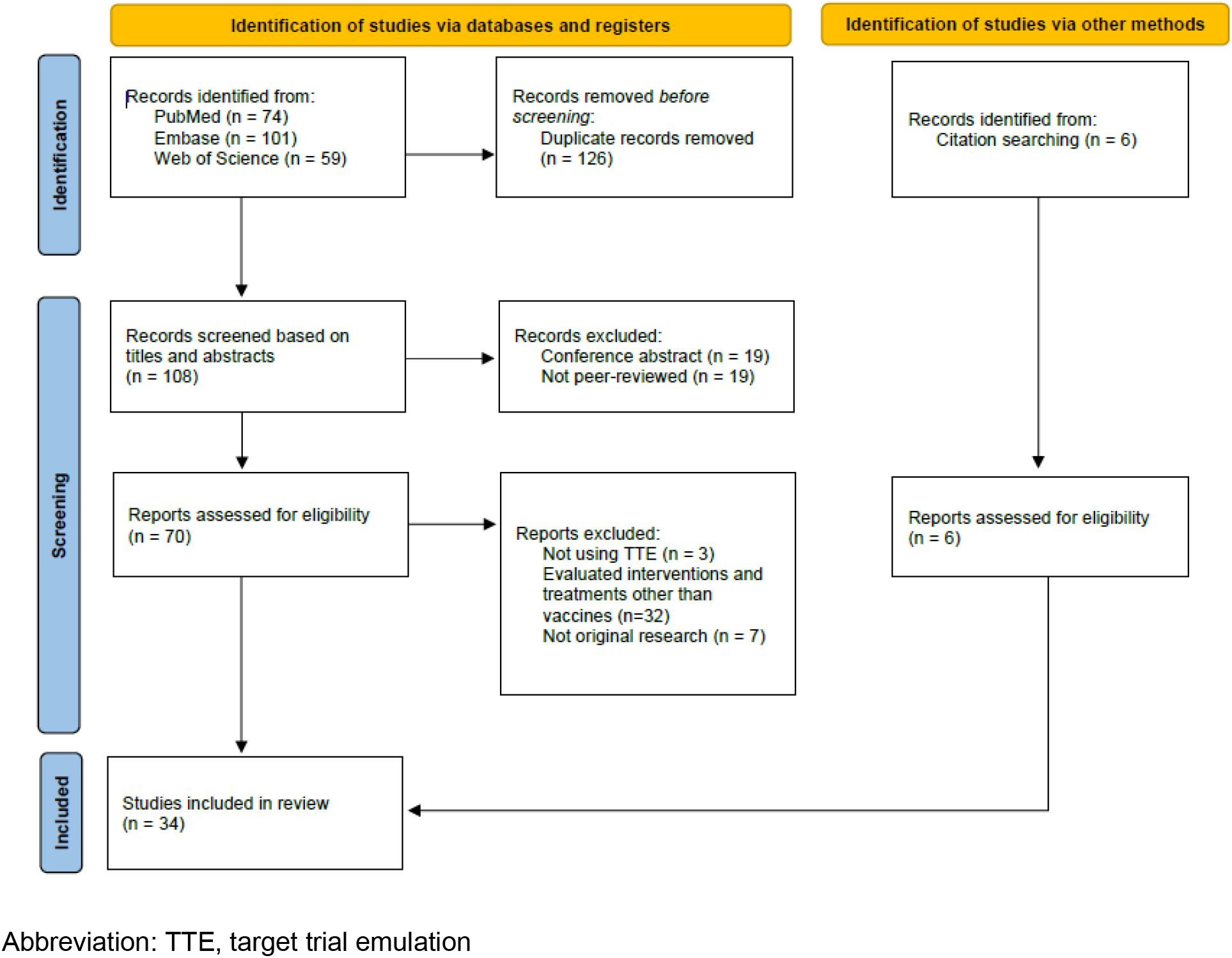
Preferred Reporting Items for Systematic Reviews and Meta-analyses (PRISMA) 2020 flow diagram for the systematic review.

**Table 1.**
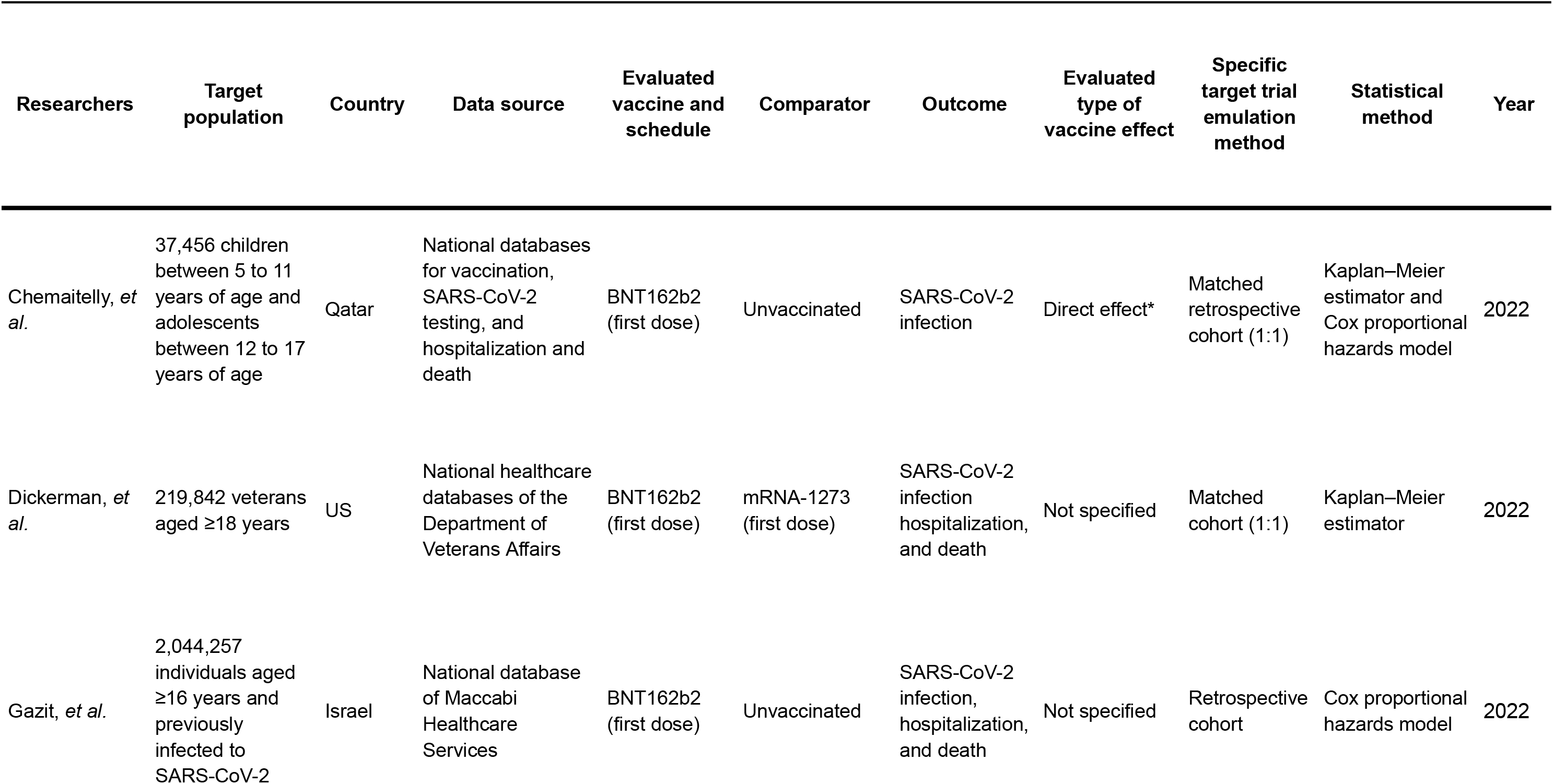
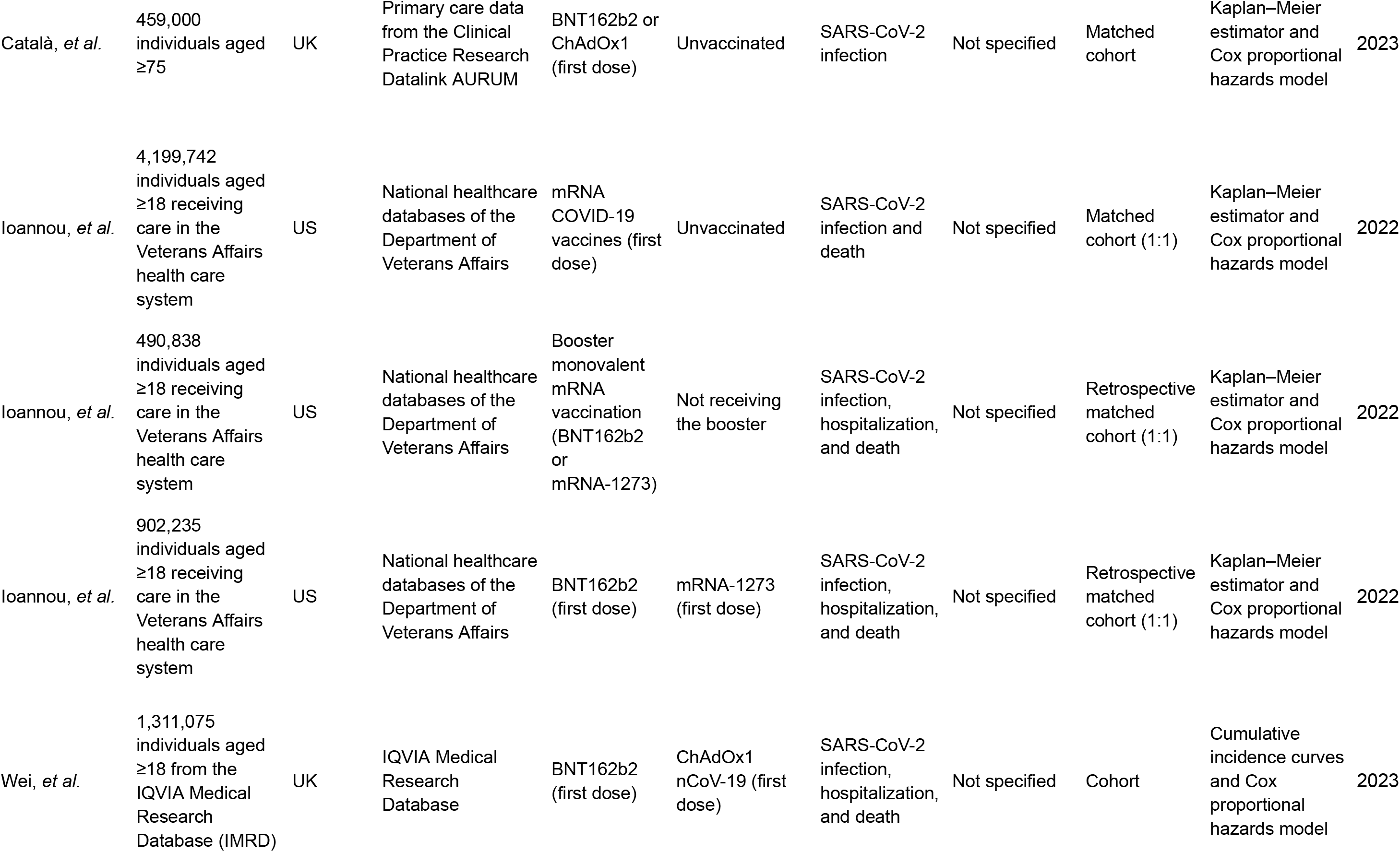
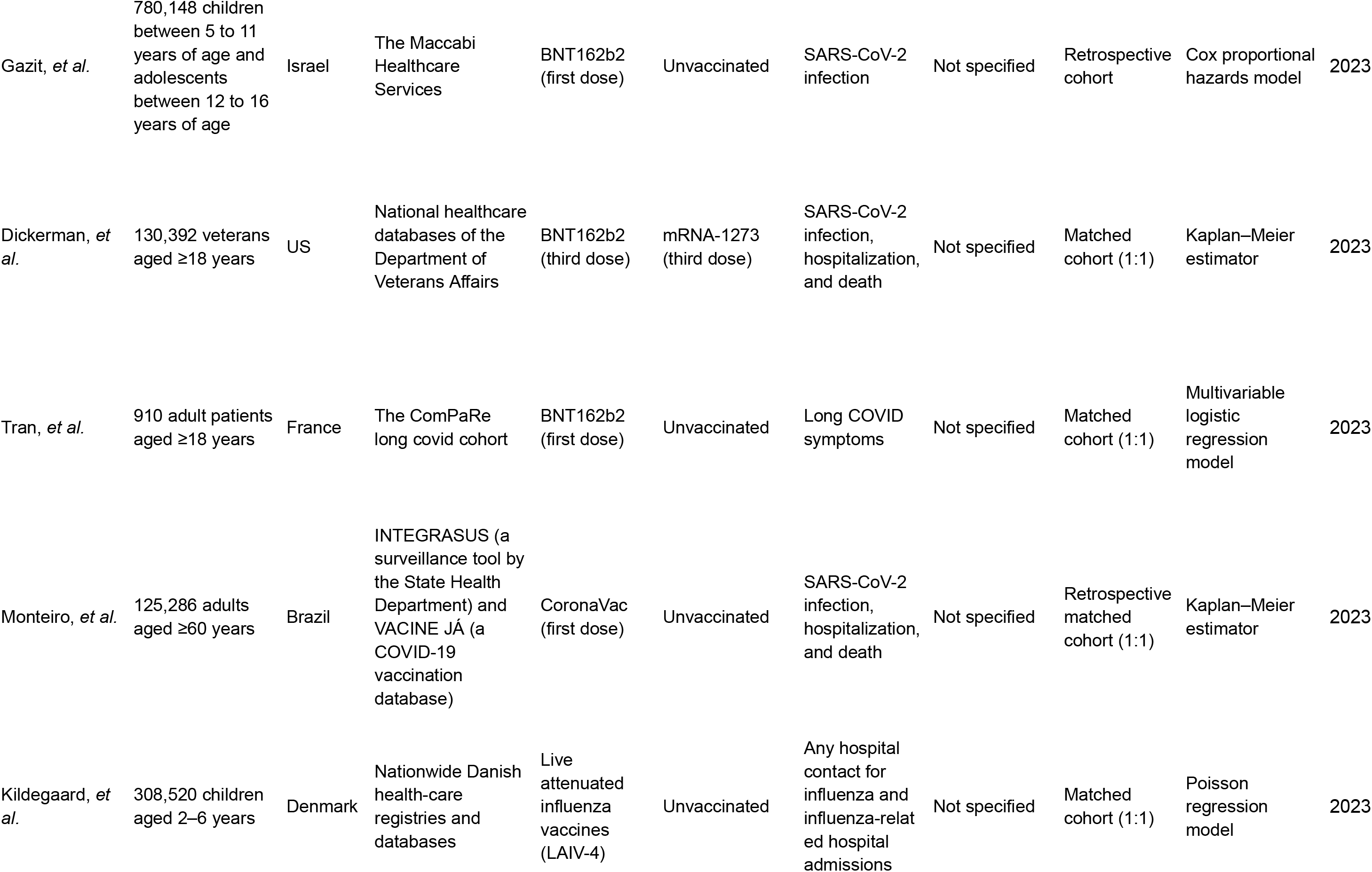
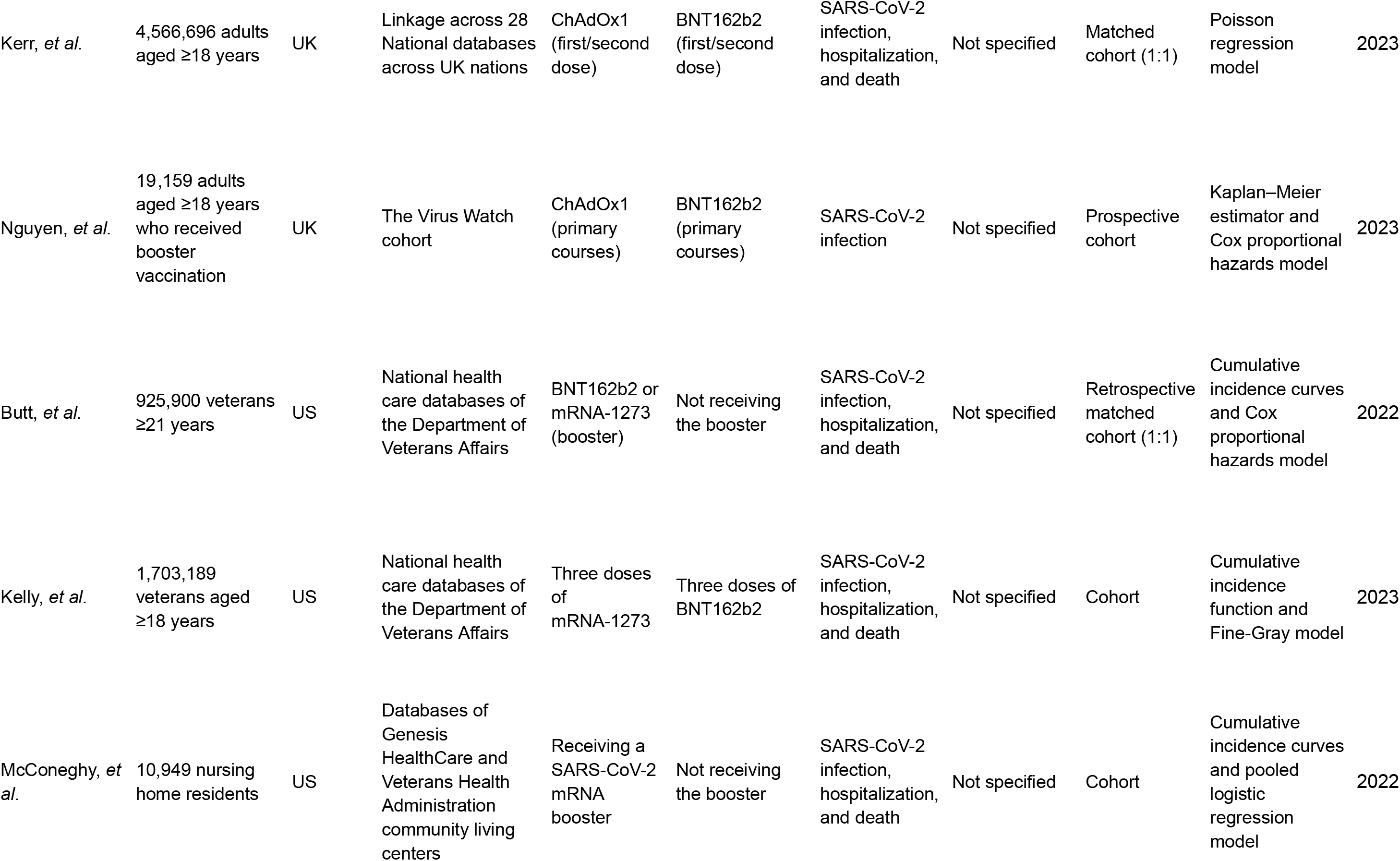
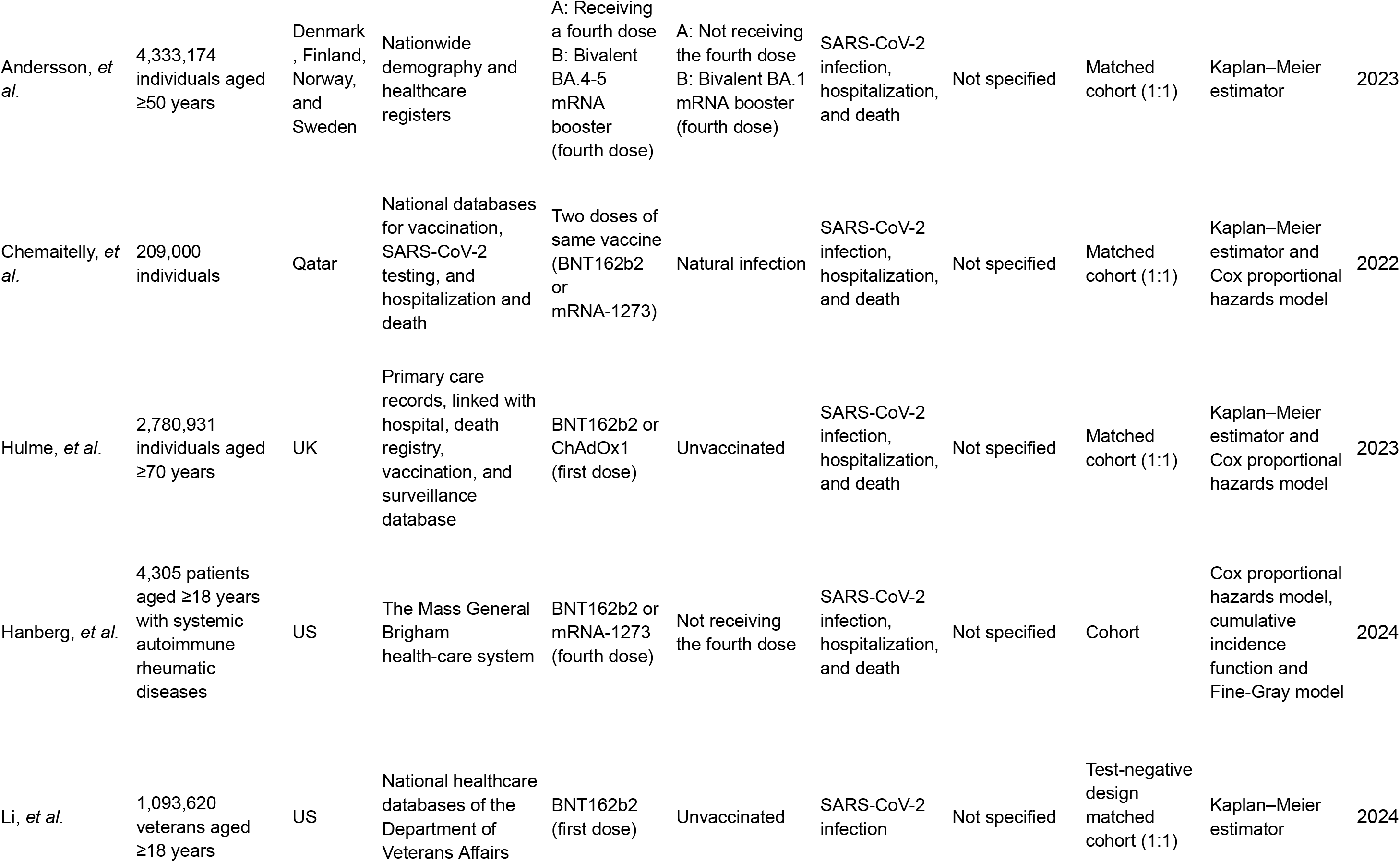
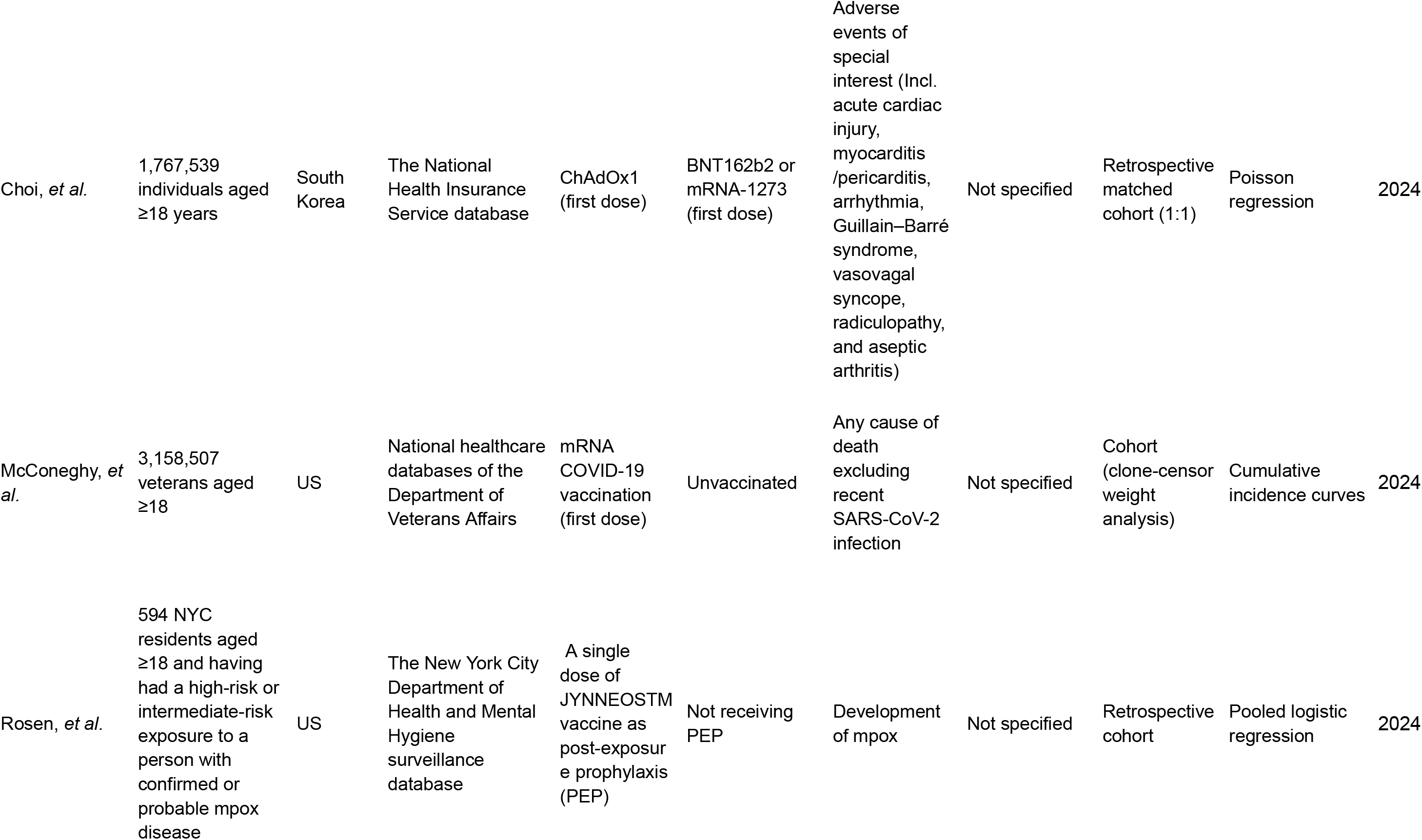
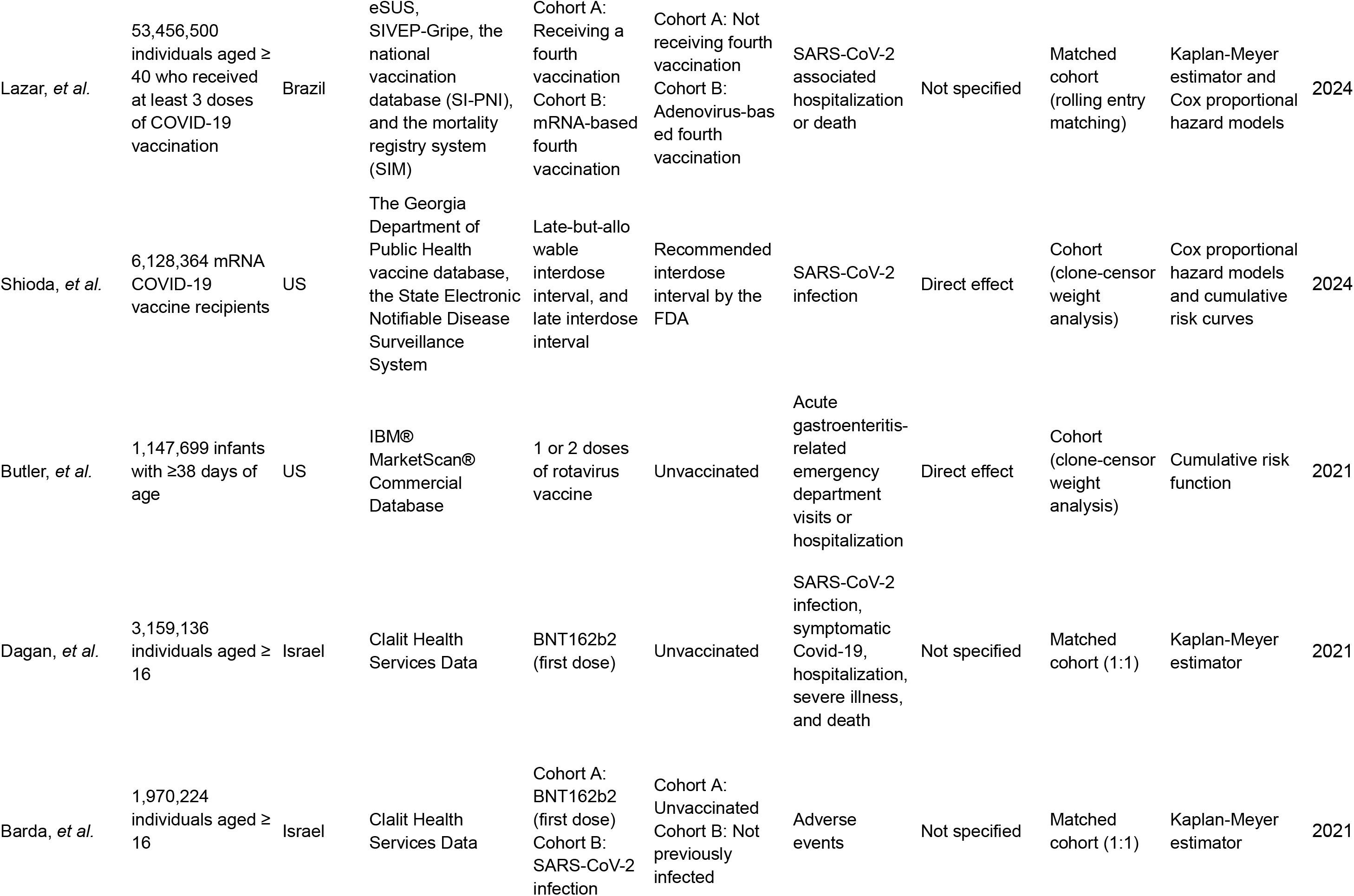
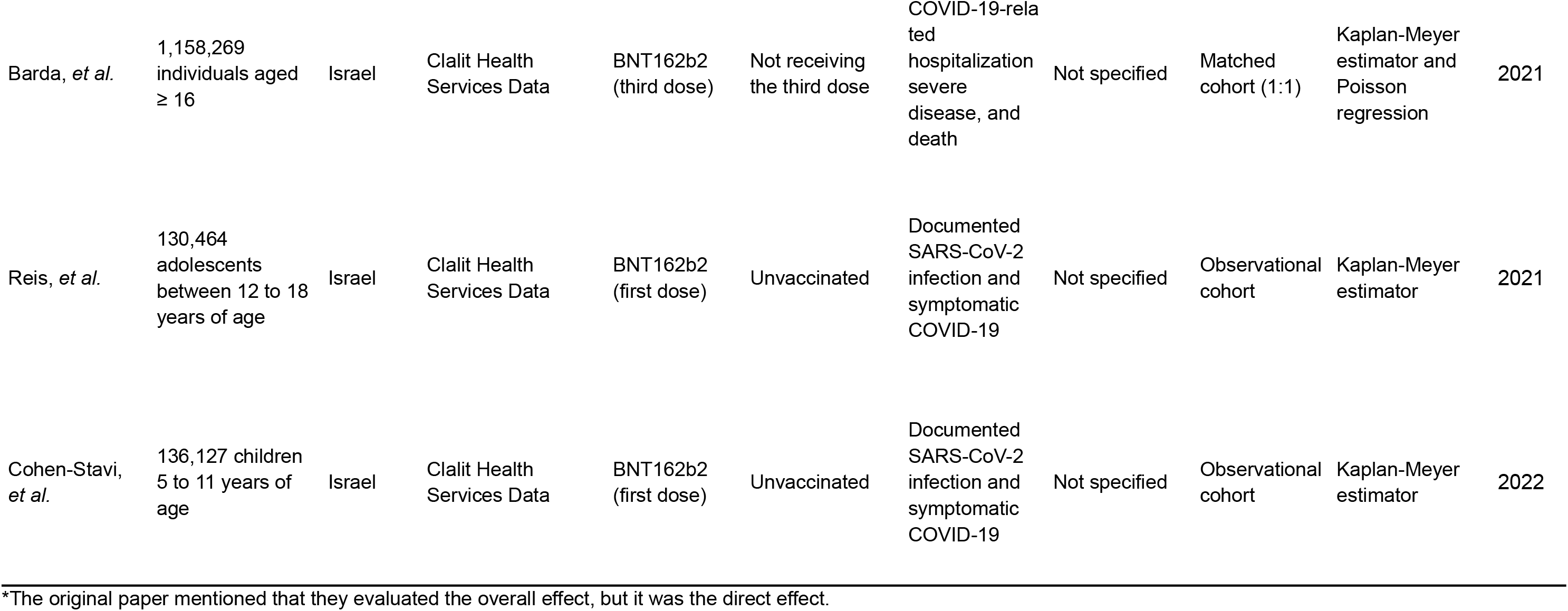
Characteristics of included studies.

### Evaluated vaccines, outcomes, and comparators

Almost all studies (n= 31, 91%) evaluated COVID-19 vaccines (23–34,36–47,49,50,52–56), with one study each evaluating influenza, mpox, and rotavirus vaccines (35,48,51). Among the COVID-19 vaccine studies, 18 (53%) examined the effectiveness against hospitalization and death (24,25,28–30,32,34,36,38–44,49,52,54), eight (24%) evaluated the effectiveness against infection only (23,26,31,37,45,50,55,56), one (3%) evaluated the effectiveness against death only (47), and one (3%) evaluated the effectiveness against infection and death but not hospitalization (27). One study (3%) focused on long COVID symptoms (46), and two studies (6%) evaluated the safety of COVID-19 vaccines against adverse events (46,53). Fourteen studies (41%) compared outcomes among vaccinated and unvaccinated individuals (23,25–27,31,33,34,43,45,47,52,53,55,56), while seven studies (21%) compared outcomes among those who received a booster dose and those who did not (28,38,40,41,44,49,54). Ten studies (29%) compared outcomes among those who received two different types of COVID-19 vaccines, such as BNT162b2 and mRNA-1273 (24,29,30,32,36,37,39,41,46,49). Two studies (6%) used individuals who had previous infections as a comparator (42,53), and one study (3%) evaluated the comparative effectiveness of different dosing intervals (50).

### TTE approaches and statistical methods

Twenty one studies (62%) used a matched cohort (23,24,26–29,32–36,38,41–43,45,46,49,52–54), while three studies (9%) used the clone-censor weight analysis to emulate target trials (47,50,51). One study (3%) prospectively collected observational data to conduct TTE analysis (37). Various statistical methods were used to quantify and compare vaccine effectiveness and safety across comparators. The majority (n= 25, 74%) used the Kaplan-Meier estimator, Cox proportional hazards model, or both (23–32,34,37,38,41–45,49,50,52–56). Seven studies (21%) applied the cumulative incidence function, the Fine-Gray subdistribution hazards model, or both to explicitly account for competing risks (30,38–40,44,47,51). Four studies (12%) used Poisson regression (35,36,46,54). Five studies (15%) applied a causal survival analysis framework to evaluate vaccine effectiveness (33,40,47,48,51).

### Evaluated types of vaccine effects and interference

Among the included studies, three studies (9%) defined the type of vaccine effect that they evaluated, all of which were direct effects (23,50,51). We did not find any study that explicitly incorporated indirect effects in vaccine evaluation. Only three studies (9%) mentioned the lack of adjustment for interference as a limitation (40,50,51).

## DISCUSSION

Our review showed that TTE approaches have increasingly become a popular method for vaccine evaluation in recent years, particularly following the introduction of COVID-19 vaccines. The majority of the included studies focused on the evaluation of COVID-19 vaccines, but TTE started to be applied to other vaccines, such as rotavirus vaccines, influenza vaccines, and mpox vaccines. Many studies did not specify which type of vaccine effects – direct, indirect, total, or overall effect – they evaluated. Interference of infectious diseases was not incorporated or addressed in vaccine evaluation in the included studies.

TTE approaches prove useful to evaluate the comparative effectiveness and safety of different vaccines (e.g., BNT162b2 and mRNA-1273 for COVID-19) or various dosing schedules (e.g., single dose vs. two doses of rotavirus vaccines; with vs. without a booster dose of COVID-19 vaccines), using observational data. RCTs cannot practically evaluate the full spectrum of dosing schedules (combination of a total number of doses and timing of each dose) in fine scale due to resource constraints. Real-world observational data, in contrast, have great variations in actual dosing schedules followed by vaccine recipients, which could be used in TTE approaches for evaluation. Selection bias and immortal time bias are major concerns when using observational data for intervention evaluation, as observational studies inherently assume that individuals successfully completed their assigned protocols, with no dropouts or outcomes during the course of intervention. This assumption is frequently violated when evaluating vaccines, especially for those requiring multiple doses, because vaccine schedules themselves affect the completion rate of the schedules. For example, the completion rate would be lower for schedules with a larger number of doses (e.g., with vs. without booster dose). Clone-censor weight analysis was specifically designed to address these issues by aligning the beginning of the follow-up time period with the date when individuals became eligible for and assigned to each protocol, rather than the date when they completed each protocol (58). These modern techniques are a promising tool to evaluate the impact of vaccines, especially when RCTs are not feasible or suitable.

Many of the included studies evaluated vaccines among the general population. This is a significant advantage of TTE approaches using large, routinely collectedd data, compared to RCTs that generally focus on smaller populations in selected settings. However, global disparity remained in the TTE studies. Most studies in our scoping review were from the U.S. and Europe. Since the impact of vaccines can vary across different settings due to varying population characteristics, disease burden, and transmission levels, data from diverse populations and regions are critical to guide global and local vaccine policy. TTE could be an affordable tool to achieve this if countries have surveillance databases and vaccine registry databases (3). To facilitate the use of TTE, it is important to teach these methods in workshops or include them in public health training curricula worldwide.

Despite various advantages of TTE, challenges persist in the application of TTE to vaccine evaluation, particularly concerning interference. The assumption of no interference is violated in studies of vaccines (59,60), because vaccines reduce population-level transmission and subsequently lower the risk of infection among both vaccinated and unvaccinated individuals.

As different vaccine products and their dosing schedules may produce varying levels of indirect effects, it is crucial to incorporate these effects into vaccine evaluation to identify optimal vaccine strategies. None of the studies in our scoping review addressed or incorporated interference in their vaccine evaluation, although a few mentioned it as a limitation. A few studies specified the types of vaccine effects evaluated, all of which were the direct effect.

Further discussions are needed to establish formal TTE methods for estimating the four effects of vaccination in the presence of interference, using large, routinely collected data. Halloran and Hudgens have discussed ways to estimate these four effects of vaccination with big data, addressing specific data requirements (61). Perez-Heydrich, *et al*. have estimated the four effects of cholera vaccination by applying inverse-probability weighted estimators to an individually-randomized controlled trial (62,63). This approach could potentially be extended to the TTE methodology. Additionally, agent-based modeling or other types of simulation modeling could be alternative approaches to addressing interference within the TTE framework. For example, agent-based modeling has been used to investigate the causal effects of preexposure prophylaxis among men who have sex with men in the presence of interference or spillover (64,65). The issue of interference is not restricted to infectious disease research; it is also relevant in fields such as health promotion programs, social epidemiology, mental health, and drug use disorder (66–70). Further work is needed to explore methods that address interference in various contexts of the target trial emulation framework.

The reliance on conventional survival analysis methods like Kaplan-Meier curves and Cox proportional hazards models in TTE studies raises concerns regarding potential biases, especially concerning competing risks (71). This is of particular concern when using surveillance data in which studies generally assume that if individuals do not show up in surveillance databases they did not have an outcome. To enhance the robustness of vaccine evaluation, we advocate for the adoption of causal survival analysis frameworks, such as pooled logistic regression with inverse probability weighting, which mitigate bias in long-term follow-up settings (72). Novel measures like restricted mean survival time (RMST) offer alternatives to evaluate vaccine effectiveness on an absolute scale, complementing traditional hazard ratio estimations (73).

A limitation of our study is the potential omission of studies that applied TTE approaches but did not explicitly label them as such. As TTE methodologies are relatively new, with standardized protocols recently being proposed and implemented (22), our keyword-based search might not have captured all relevant studies. To mitigate this issue, we conducted a review of the literature and examined the citations of identified studies to include any that were initially missed in our keyword-based search. Nonetheless, this approach may not have been exhaustive.

In summary, TTE has various practical and methodological advantages and is a useful approach for evaluating vaccine programs, especially when RCTs are not feasible. Clone-censor weight analysis is particularly effective for evaluating multi-dose vaccines to avoid selection bias and immortal time bias. Challenges related to interference need to be addressed to incorporate indirect effects or spillover effects in the evaluation of interventions, including vaccines. Moving forward, further collaboration between causal inference researchers and infectious disease experts will be beneficial for developing new approaches and guidelines on the application of TTE for vaccine evaluation in the presence of interference.

## Data Availability

All data produced in the present study are available upon reasonable request to the authors

## ACKNOWLEDGEMENT

We thank Kristen Sheridan for her support with the literature review. We also thank Dr. M. Elizabeth Halloran, Dr. Marc Lipsitch, and Katherine Jia for their invaluable feedback. This study was also made possible by cooperative agreement CDC-RFA-FT-23-0069 from the CDC’s Center for Forecasting and Outbreak Analytics. Its contents are solely the responsibility of the authors and do not necessarily represent the official views of the Centers for Disease Control and Prevention.

---

**Supplementary Table 1.** Search terms and results of the literature review.

| Search word | Medium | Start Date | End Date | Result |
| --- | --- | --- | --- | --- |
| ((("2012/01/01"[Date - Publication] : "2024/5/21"[Date - Publication])) AND (((target trial[Title/Abstract]) OR (target emulation[Title/Abstract]) OR (trial emulation[Title/Abstract]) OR (targeted trial[Title/Abstract]) OR (emulated trial[Title/Abstract]) OR (emulated target[Title/Abstract])) AND ((infectious diseases[Title/Abstract]) OR (infection[Title/Abstract]) OR (vaccine[Title/Abstract]) OR (vaccination[Title/Abstract]) OR (immunization[Title/Abstract])))) | PubMed | 2012/01/01 | 2024/05/21 | 74 |
| ('target trial':ti,ab OR 'target emulation':ti,ab OR 'trial emulation':ti,ab OR 'targeted trial':ti,ab OR 'emulated trial':ti,ab OR 'emulated target':ti,ab) AND ('infectious diseases':ti,ab OR 'infection':ti,ab OR 'vaccine':ti,ab OR 'vaccination':ti,ab OR 'immunization':ti,ab) | Embase | 2012/01/01 | 2024/05/21 | 101 |
| (TI=("target trial" OR "target emulation" OR "trial emulation" OR "targeted trial" OR "emulated trial" OR "emulated target")OR AB=("target trial" OR "target emulation" OR "trial emulation" OR "targeted trial" OR "emulated trial" OR "emulated target")) AND (TI=("infectious diseases" OR "infection" OR "vaccine" OR "vaccination" OR "immunization") OR AB=("infectious diseases" OR "infection" OR "vaccine" OR "vaccination" OR "immunization")) | Web of Science | 2012/01/01 | 2024/05/21 | 59 |

\*Results as of May 21, 2024

